# A Cost-utility Analysis of Ferric Derisomaltose versus Ferric Carboxymaltose in Patients with Iron Deficiency Anemia in China

**DOI:** 10.1101/2024.07.11.24310267

**Authors:** F Zhang, A Shen, Waqas Ahmed, Richard F. Pollock

## Abstract

**Aims:** Intravenous (IV) iron is the recommended treatment for patients with iron deficiency anemia (IDA) who are unresponsive to oral iron treatment or require rapid iron replenishment. Ferric derisomaltose (FDI) and ferric carboxymaltose (FCM) are high-dose, rapid infusion, IV iron formulations that have recently been compared in three head-to-head randomized controlled trials (RCTs), which showed significantly higher incidence of hypophosphatemia after administration of FCM than FDI. The present study objective was to evaluate the cost-utility of FDI versus FCM in a population of Chinese patients with IDA.

**Materials and methods:** A previously-published patient-level simulation model was used to model the cost-utility of FDI versus FCM in China. The number of infusions of FDI and FCM was modeled based on the approved posology of the respective formulations using simplified tables of iron need in a population of patients with bodyweight and hemoglobin levels informed by a Chinese RCT of FCM. Data on the incidence of hypophosphatemia was obtained from the PHOSPHARE-IDA RCT, while data on disease-related quality of life was obtained from SF-36v2 data from the PHOSPHARE-IBD RCT.

**Results:** Over the five-year time horizon, patients received 3.98 courses of iron treatment on average, requiring 0.90 fewer infusions of FDI than FCM (7.69 versus 6.79). This resulted in iron procurement and administration cost savings of RMB 206 with FDI (RMB 3,519 versus RMB 3,312). Reduced incidence of hypophosphatemia resulted in an increase of 0.07 quality-adjusted life years and further cost savings of RMB 782 over five years, driven by reduced need for phosphate testing and replenishment. FDI was therefore the dominant intervention.

**Conclusions:** Results showed that FDI would improve patient quality of life and reduce direct healthcare expenditure versus FCM in patients with IDA in China.

## Introduction

One of the most common causes of anemia is iron deficiency (ID) [1]. Iron deficiency anemia (IDA) affects 30% of the worldwide population, is the most common nutritional deficiency in the world, and is a significant contributor to the global disease burden [2–4]. In China IDA is estimated to affect 176 million people [5], and 13.9% of pregnant women [6]. This already substantial IDA burden is likely to increase, based on the fact that childhood obesity prevalence is rising in China [7] and children with obesity have been found to possess a relatively higher risk of developing ID and IDA [8,9].

Some factors that have been linked to causing IDA include menstruation in women, gastrointestinal bleeding, decreased dietary iron intake, and absorption issues [10]. Symptoms of IDA can vary from patient to patient, but may include shortness of breath, palpitations, fatigue and tachycardia due to lower blood oxygen levels, with more serious manifestations including angina, abdominal pain and motility disorders [2]. Evidence from the published literature demonstrates that IDA can significantly impact the quality of life (QoL) of those living with the condition, particularly pregnant or heavily menstruating women [11–13], individuals with chronic heart failure [14] and those with inflammatory bowel disease (IBD) [15]. Despite these impacts, however, treating individuals living with IDA has been shown to lead to QoL improvements, regardless of the underlying cause of the condition [16,17].

Treatment for IDA conventionally involves the use of oral iron supplementation, which are well responded to by most patients [18]. However, in patients where oral iron is contraindicated, does not allow for rapid anemia correction, or is indeed ineffective (i.e., those with IBD) [19,20], intravenous (IV) iron therapy is pursued, and has been demonstrated to be clinically effective [21,22].

Ferric derisomaltose (FDI) and ferric carboxymaltose (FCM) are two IV iron formulations that are used in the treatment of IDA, each allowing a large amount of iron to be administered rapidly per infusion [23], leading to faster anemia correction than oral iron. Both treatments have been demonstrated to have a comparable clinical effectiveness, but FCM is associated with significantly higher rates of hypophosphatemia [24–26]. Indeed, the PHOSPHARE-IDA randomized controlled trials (RCTs) found that 74.4% of patients receiving FCM and only 8.0% of patients receiving FDI experienced hypophosphatemia (defined as a serum phosphate level of <2.0 mg/dL) [27]. Repeated use of FCM may therefore result in hypophosphatemia-related osteomalacia, which can lead to symptoms including bone pain and fracture [23]. A further advantage of FDI relative to FCM is the higher dose per infusion that can be administered. Specifically, the maximum dose of FCM per infusion is 1000mg or iron (usually administered over 15 minutes) [28]. For FDI, elemental iron amounts administered per infusion can exceed 1000mg based on a weight-related dose of 20mg/kg [29] (although doses exceeding 1000mg would require 30 minutes or more for administration).

Cost-utility analyses are a widely-used method to determine how best to allocate scarce financial resources for healthcare interventions, by evaluating the comparative costs of using different health technologies to improve health-related outcomes [30]. Given the high prevalence of IDA in China, as well as its significant QoL and cost implications, there is a need to determine the most cost-effective IV iron replacement therapy for individuals living with IDA. The objective of the present study was therefore to evaluate the cost-utility of FDI versus FCM in patients with IDA in China.

## Methods

### Patient-level simulation model

A patient-level simulation model was developed in Microsoft Excel, based on a previously-published cost-utility model of a similar design [31]. The rationale for using a patient-level simulation model was based on the National Institute for Health and Care Excellence (NICE) Decision Support Unit’s technical guidance document [32], particularly the non-linear relationship between patient baseline characteristics (e.g., hemoglobin levels and bodyweight) and intermediate model outcomes (e.g., iron need). Other facets of the analysis that informed the decision to utilise a patient-level simulation model were the need to incorporate parameter-level stochastic variation, stochastic uncertainty and individual patient variability.

Each simulated patient was assigned baseline values for age, bodyweight, and hemoglobin randomly sampled from baseline distributions. The mean iron need for each patient was then determined using these assigned baseline values combined with the simplified tables of iron need from the respective summaries of product characteristics (**Table 1**), alongside the total number of treatment courses that each patient would require over one complete model cycle.

**Table 1.** Simplified tables of iron need from the Chinese summaries of product characteristics for ferric carboxymaltose and ferric derisomaltose.

| Ferric carboxymaltose |  |  |  |  |
| --- | --- | --- | --- | --- |
| Hemoglobin |  | Bodyweight |  |  |
| g/dL | mmol/L | <35 kg | ≥35 kg and <70 kg | ≥70 kg |
| <10 | <6.2 | 500 mg | 1,500 mg | 2,000 mg |
| ≥10 and <14 | ≥6.2 and <8.7 | 500 mg | 1,000 mg | 1,500 mg |
| ≥14 | ≥8.7 | 500 mg | 500 mg | 500 mg |
| Ferric derisomaltose |  |  |  |  |
| g/dL | mmol/L | <50 kg | ≥50 kg and <70 kg | ≥70 kg |
| <10 | <6.2 | 500 mg | 1500 mg | 2000 mg |
| ≥10 | ≥6.2 | 500 mg | 1000 mg | 1500 mg |

Based on the modeled number of infusions administered per cycle, QoL process-related disutilities (decrements in QoL associated with patients engaging with a particular treatment process) and costs associated with the administration of IV iron were calculated. An equal proportion of patients in both treatment arms were assumed to achieve hematological response, stemming from PHOSPHARE-IBD RCT results [33].

PHOSPHARE-IBD RCT results were also used as the basis of QoL data for disease-related events specific to treatment. Monte Carlo methods were employed to explore alternative results and outcomes for simulated patients, using the built-in individual-level variability and parameter-level stochastic variation. Comprehensive QoL outcomes and costs over the complete time horizon were determined for each patient in each model cycle. Results were recorded and analyzed as part of the entire patient cohort results dataset.

Cost outcomes were reported in 2023 Renminbi (RMB) and were recorded and summarized alongside quality-adjusted life year (QALY) and survival outcomes. The final incremental cost-utility ratio (ICUR) was calculated using the differences in these outcomes between the FDI and FCM arms.

### Clinical data and patient characteristics

A pragmatic search of the published literature was conducted to identify data on the safety and effectiveness of FDI and FCM. Specifically, a 2019 systematic review [34] and 2020 network meta-analysis investigating IV-iron related hypophosphatemia [35] were used to identify relevant studies. Data on hypophosphatemia incidence and the safety of the two IV iron formulations were also obtained from two additional literature reviews [24,36]. To ensure that all clinical information was up-to-date, a further supplementary search was conducted to identify relevant RCTs published after the original pragmatic search was completed.

The model assumed that both FDI and FCM treatment would result in the same time to recurrence of anemia, and an equivalent initial increase in hemoglobin levels. This assumption was ultimately based on the findings of the PHOSPHARE-IBD RCT, which reported no significant difference in hematological response between the FDI and FCM arms.

Chinese life tables were used to model background mortality. Simulated baseline patient characteristics were obtained from the NCT03591406 RCT, which investigated FCM compared with iron sucrose in patients with IDA in China [37]. These characteristics included bodyweight (60.07 kg), mean age (39.4 years), and pre-treatment hemoglobin levels (7.9 g/dL; **Table 2**). The median time to iron retreatment was taken to be 16 months [38]. Regarding adverse effects, 3.4% of patients receiving FDI and 65.1% of patients receiving FCM experienced any hypophosphatemia, [27]. Hypophosphatemic osteomalacia was conservatively omitted from the analysis. Severe hypophosphatemia was modeled in 0.0% of patients receiving FDI, and 7.3% of patients receiving FCM, based on results from Wolf et al. [27].

**Table 2.** Baseline patient characteristics, median time to retreatment, and hypophosphatemia incidence rates.

| Item | Baseline value |  | Source |
| --- | --- | --- | --- |
| Baseline mean (SD) age, years | 39.40 (9.31) |  | NCT03591406 [37] |
| Baseline mean (SD) bodyweight, kg | 60.07 (12.0) |  |  |
| Pre-treatment mean (SD) hemoglobin level, g/dL | 7.9 (1.48) |  |  |
| Median time to IDA recurrence, months | 16.00 (4.34) |  | Kulnigg et al.[38] |
| **Hypophosphatemia** | **FDI** | **FCM** |  |
| Patients experiencing any hypophosphatemia (<2mg/dL) % | 3.4 | 65.10 | Zoller et al.[33] |

**Abbreviations:** FCM, ferric carboxymaltose; FDI, ferric derisomaltose; IDA, iron deficiency anemia; SD, standard deviation.

### Perspective, time horizon, and discounting

Costs were investigated from the perspective of the Chinese healthcare system. All costs were calculated and reported in 2023 RMB. An annual discount rate of 5.0% was applied to future QoL effects and costs, based on 2020 guidelines for pharmacoeconomic evaluations in China [39]. As no formal willingness-to-pay (WTP) threshold is currently published in China, a WTP threshold of 85,698 RMB was used, representing the 2022 Chinese Gross Domestic Product (GDP) *per capita* [40].

A five-year time horizon was adopted to ensure that the chronic and long-term impacts of IDA and its treatment could be effectively captured, by ensuring that multiple IDA treatment cycles would be included within the analysis. A monthly cycle length was adopted in the model, allowing iron treatment courses and hypophosphatemia-related monitoring and treatment to be captured over multiple intra-annual cycles to better reflect real-world practice. This also allowed a gradual reversion to baseline hemoglobin levels in patients with chronic underlying etiologies of IDA to occur within a single year, whilst alleviating computational burden relative to shorter cycle lengths.

### Resource use

For each patient receiving FCM, it was assumed that two phosphate tests would be performed during each treatment course, based on recommendations from the published literature [41], a 2020 Drug Safety Update from the UK Medicines and Healthcare products Regulatory Agency (MHRA) focused specifically on hypophosphatemia (and subsequent osteomalacia events) associated with FCM [42], and the Chinese summary of product characteristics for FCM [43]. It was assumed that no serum phosphate tests would be conducted in patients treated with FDI, and that no patients would therefore receive either phosphate tests or treatment with exogenous phosphate. This assumption was informed by the absence of any severe (and therefore more likely symptomatic) hypophosphatemia in patients treated with FDI in the PHOSPHARE-IBD RCT. Conversely, with FCM, it was assumed that all patients in whom moderate or severe hypophosphatemia was present (in line with the incidence rates from the PHOSPHARE-IBD RCT) would receive IV phosphate therapy in line with recommendations from clinicians in China. For patients experiencing moderate hypophosphatemia, four phosphate infusions were assumed to be required for treatment, with six phosphate infusions assumed for patients experiencing severe hypophosphatemia [44].

### Health-related quality of life data

Health state utility values were derived from the SF-36v2 generic patient-reported outcome questionnaire as administered in the PHOSPHARE-IBD RCT [45]. Participants in the trial were subject to identical dosage regimens based on their weight and hemoglobin levels, and received either FDI or FCM at two defined time intervals (baseline and day 35). SF-6Dv2 utility values were used as the basis for calculating differences in QoL for FDI and FCM recipients. To determine the SF-6Dv2 health state utility values (HSUVs), the study used ten items from the SF-36v2 questionnaire. The ten items were combined to form six health dimensions, each comprising 5-6 distinct health levels. To calculate the final SF-6Dv2 HSUVs, utility weights were applied, before summing the weighted scores for each of the six dimensions. Each weight comprised a negative value and would result in a decrement from the initial utility score of 1 (which represents full health). However, if the worst score for any dimension was experienced by patients, then an additional utility decrement was also applied [41]. The study applied China-specific process-disutilities (decrements in QoL associated with patients engaging with or experiencing a particular treatment process, rather than due to the disease itself) [46]. A previously-published diminishing marginal utility model was incorporated for this purpose, and determined QoL associated with IV iron infusions [47]. Specifically, as patients received more IV iron infusions, they would experience a marginal disutility that decreased on a logarithmic scale as the total infusions they received increased. The concept for this diminishing marginal disutility was to convey how patients may adapt to receiving regular IV iron infusions over time, resulting in a gradually reducing treatment-related QoL burden.

### Costs

The analysis was based on a unit price of RMB 63.6 per 100mg of FDI and RMB 58.51 per 100mg pack of FCM [48,49]. The cost of administering the iron was assumed to be equal for both FDI and FCM on a per-infusion basis, at RMB 33.66 per infusion [50]. The cost of one serum phosphate test was taken to be RMB 5.00 in line with the median charge for public medical services across Beijing, Guangzhou, Shanghai, Shenzhen, and Xiamen.

Hypophosphatemia treatment costs included the cost of intravenous phosphate in addition to the costs associated with administering the infusion. Specifically, a cost of RMB 431.09 was established for six IV phosphate infusions (in cases of severe hypophosphatemia treatment), and RMB 281.28 for four IV phosphate infusions (in cases of moderate hypophosphatemia treatment) [51].

### Sensitivity analysis

A series of one-way sensitivity analyses (OWSA) was conducted to assess the likely effects of varying individual model parameter values on the base case ICUR. Discount rates, baseline bodyweight and hemoglobin levels, and intravenous phosphate costs were among the parameters varied for the OWSA. In the probabilistic sensitivity analysis (PSA), 1000 Monte Carlo iterations were conducted to explore the effect of simultaneously varying model parameters by random sampling.

### FCM price breakeven analyses

As drug prices are subject to change over time, a series of analyses were conducted in which the list price of FCM was reduced from 100% of the base case value (of RMB 292.56) to 0% of the base case interval in 10% intervals. Results of the analyses were presented in the forms of incremental costs, ICURs, and the net monetary benefit of FDI versus FCM to establish the FCM price points at which FDI and FCM would be at overall cost parity from the payer perspective, and if there were any scenarios in which the conclusions regarding the cost-effectiveness findings from the base case analysis would change as a result of the reduced price of FCM.

## Results

### Base case

Over a five-year time horizon, patients treated with FDI required 0.9 fewer iron infusions versus FCM over an average of 3.98 courses of iron (7.69 infusions with FCM versus 6.79 infusions with FDI). Differences in the simplified tables of iron need between the two IV iron formulations resulted in an average of 6,088 mg of FCM being adminstered per patient over five years, versus 5,297 mg of FDI.

The difference in the number of infusions resulted in iron procurement and administration cost savings of RMB 206 (RMB 3,519 for FCM versus RMB 3,312 for FDI) and a process utility-driven gain of 0.0042 QALYs with FDI versus FCM. FDI was also associated with an additional 0.0663 QALYs based on hypophosphatemia-related fatigue as reported in the PHOSPHARE-IBD SF-6D data, and further cost savings of RMB 782, driven by the absence of serum phosphate testing and administration of exogenous phosphate in patients treated with FDI. Overall, FDI therefore resulted in cost savings of RMB 989 and an improvement in quality-adjusted life expectancy of 0.0705 QALYs over five years. FDI was therefore the dominant intervention, associated with increased clinical benefit at a lower cost than FCM. A summary of the base case results is presented in **Table 3**.

**Table 3.** Health economic results from the base case analysis.

|  | **FCM** | **FDI** | **Incremental** |
| --- | --- | --- | --- |
| **Base case (Chinese healthcare system perspective)** |  |  |  |
| **Life expectancy (years)** | **4.97** | **4.97** | **0.00** |
| **Quality-adjusted life expectancy (QALYs)** | **2.50** | **2.57** | **+0.07** |
| **Total cost (RMB)** | **4,301** | **3,312** | **-989** |
| Iron costs (RMB) | 3,280 | 3,102 | -178 |
| Iron administration costs (RMB) | 238 | 120 | -28 |
| Hypophosphatemia (RMB) | 782 | 0 | -782 |
| **Incremental cost-utility ratio** | **FDI dominant** |  |  |

Abbreviations: FCM, ferric carboxymaltose; FDI, ferric derisomaltose; QALYs, quality-adjusted life years; RMB, Renminbi.

### Probabilistic sensitivity analyses

In the PSA, ICURs from all model iterations fell within the south-eastern quadrant of the cost-utility plane (representing reduced costs and QALY-gains), with FDI consistently being the dominant intervention relative to FCM (**Figure 1**). At all WTP thresholds between RMB 0.00 and RMB 161,952, FDI was 100% likely to be a cost-effective strategy.

**Figure 1.**
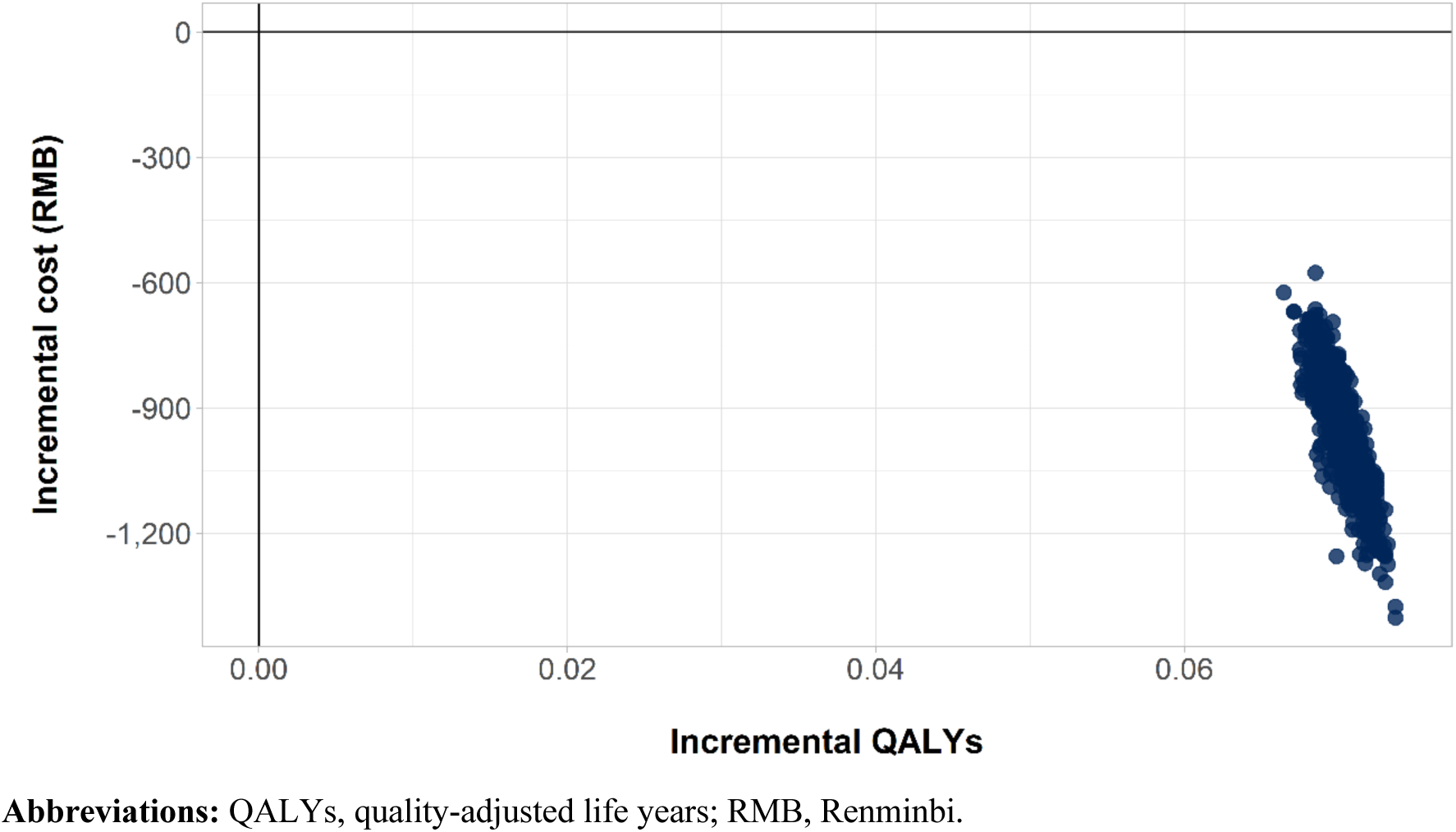
Scatterplot showing results from probabilistic sensitivity analysis on the cost-effectiveness plane.

### One-way sensitivity analysis

The ten model parameters with the largest impact on the final base case ICUR in OWSA are presented in **Figure 2**. Baseline bodyweight was the parameter with the largest effect on the ICUR. Specifically, a baseline bodyweight of 54.06kg changed the ICUR by RMB –5,649.48 per QALY gained, while a baseline bodyweight of 66.08kg resulted in a change in the ICUR of RMB +4,621.09 per QALY gained. Cost discount rate was associated with the second greatest variation in the final ICUR; a cost discount rate of 0% changed the ICUR by RMB – 1,201.69 per QALY gained, while a cost discount rate of 8% changed the ICUR by RMB +619.31 per QALY gained. Regardless of whether a higher or lower value (comparative to that from the base case analysis) was used, neither time to recurrence of IDA nor FDI cost per infusion led to any changes on the final base case ICUR.

**Figure 2.**
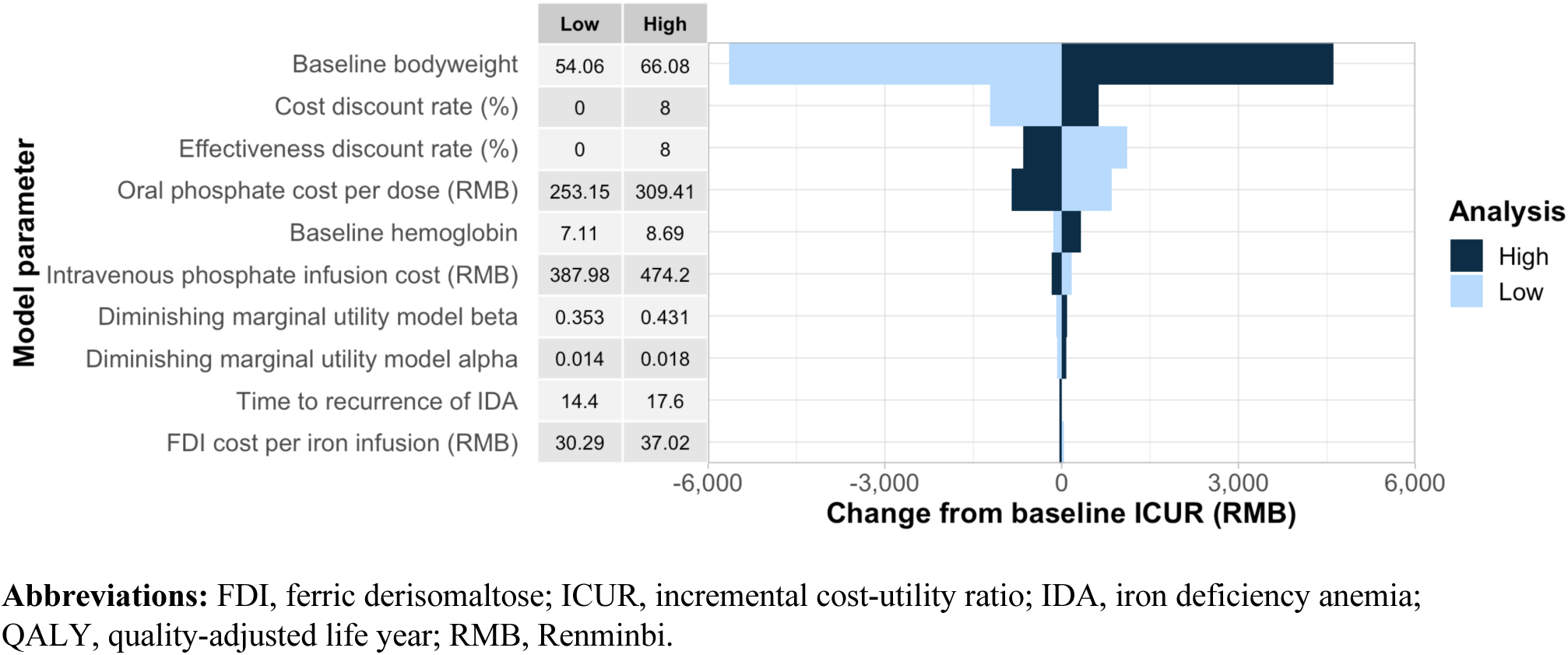
Tornado plot of results from one-way sensitivity analyses.

### FCM price breakeven analyses

Results of the FCM price breakeven analyses are presented in Figure 3. The analyses showed that FCM and FDI broke even with regard to the absolute costs from the Chinese healthcare system perspective with a 31% discount on the FCM list price used in the base case; however, there was no positive FCM price point at which FCM would be considered cost-effective relative to FDI based on a WTP threshold of RMB 85,698 per QALY, nor was there any price point at which there would be a net monetary benefit of using FCM versus FDI (Figure 3).

**Figure 3.**
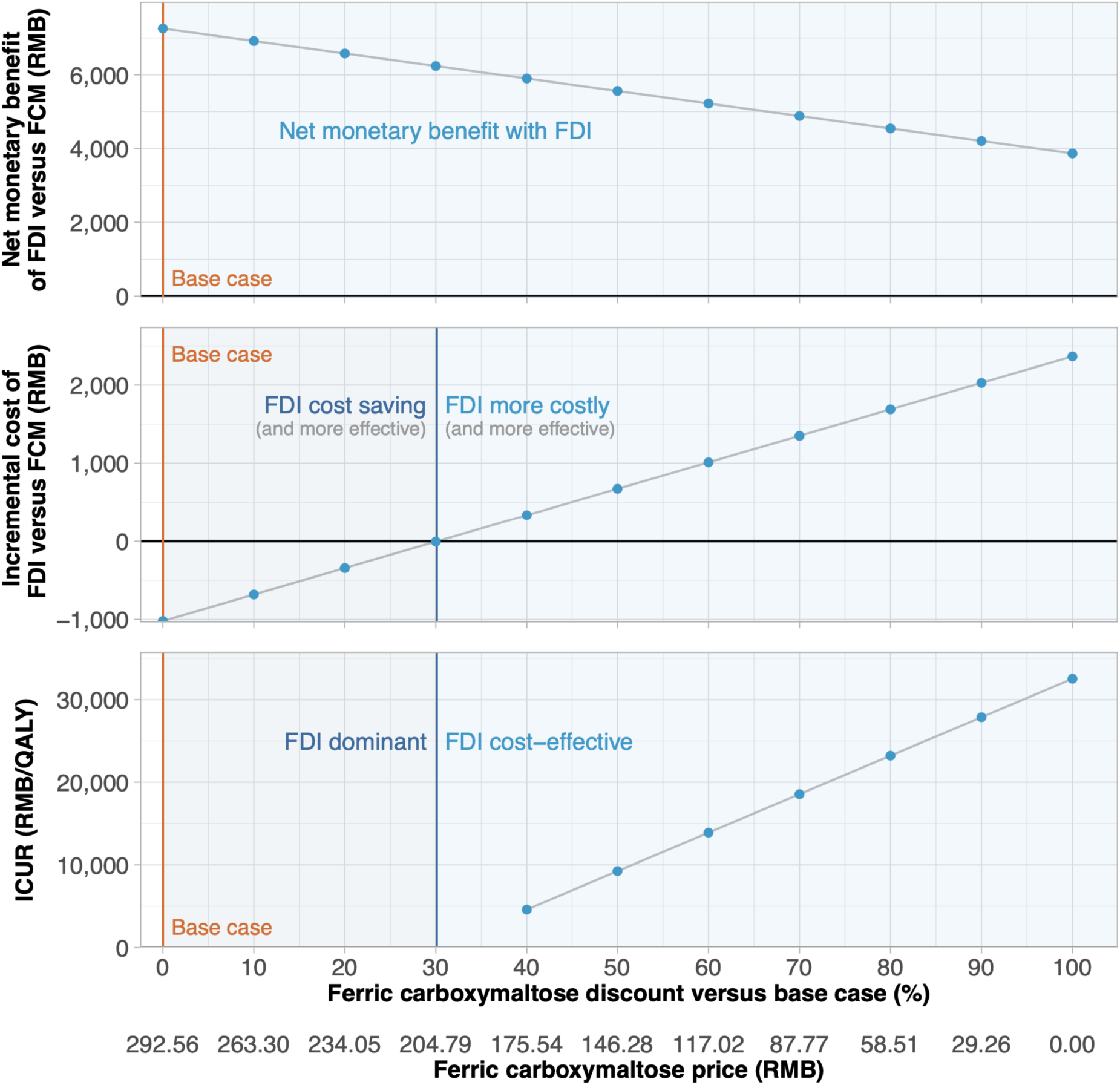
Incremental costs, incremental cost-utility ratios, and net monetary benefit outcomes from analyses in which the unit price of ferric carboxymaltose was varied from 100% to 0% of the base case value.

## Discussion

The present study determined that FDI was the dominant intervention relative to FCM and led to significantly lower incidence of hypophosphatemia. Specifically, FDI was projected to result in procurement and administration cost-savings of RMB 200 (RMB 3,512 for FCM versus RMB 3,312 for FDI) and 0.07 QALY gains (2.50 QALYs for FCM and 2.57 QALYs for FDI) over a five-year time horizon. An additional cost difference of RMB 782 stemmed from hypophosphatemia monitoring and treatment (RMB 782 for FCM, and RMB 0.0 for FDI). Reduced iron administration costs for FDI were driven by patients receiving FCM requiring 0.9 additional infusions over a five-year time horizon compared to patients receiving FDI. The additional 0.9 infusions for FCM treatment may also be associated with increased productivity losses (specifically absenteeism) due to hospital visits and infusion times. However, these costs were not incorporated into the present model. Similarly, cardiovascular (CV) and hypersensitivity (HSR) event incidence (and subsequent financial burden) were also omitted from the model, with their inclusion likely to demonstrate FDI to be even more cost-effective than the current results suggest. This is because FCM is associated with comparatively higher CV and HSR even incidence than FDI [52,53].

To the knowledge of the authors, this cost-utility study of FCM versus FDI in patients with IDA is the first to be conducted within a Chinese healthcare setting. However, the final results were aligned with those from similar studies conducted within European settings, which also found FDI to be cost-effective relative to FCM. In the European studies, additional costs associated with FCM primarily stemmed from increased infusion numbers [54,55]. The collective evidence suggests that FDI is likely to be cost-effective compared to FCM across a wide range of countries and healthcare settings, even when modelled patient characteristics in terms of hemoglobin levels, co-morbidities and weight vary geographically.

A strength of this study was the use of China-specific sources to gather relevant model inputs. Specifically, costs associated with hospital stays, hypophosphatemia treatment, IV iron infusions, and serum phosphate tests were all obtained from sources specific to China. With regard to utility values, the disutilities associated with infusions were obtained from a time trade-off valuation study conducted with Chinese patient participants. In cases where data specific to China were unavailable, efforts were made to identify suitable proxies. Disease-related health state utility values were derived from the PHOSPHARE-IBD RCT, which enrolled a subpopulation of people living with IDA (i.e., those also living with IBD) and was conducted across five European countries [45]. When interpreting the results of the present study, the European provenance of the QoL data, and the etiology of the IDA (i.e., IBD) should be considered; populations with different underlying etiologies of IDA in different geographies may experience different QoL effects versus a general IDA population in China. For instance, fatigue is a known driver of reduced QoL in patients with IBD and IDA [56] and has been shown to be exacerbated by hypophosphatemia; however, it may not rank among the largest drivers of reduced QoL in a population of patients with IBD who may also experience symptoms such as diarrhea and constipation, abdominal pain, bleeding or abnormal discharge [57]. In patients who experience IDA as a consequence of other conditions (where the range of possible symptoms are narrower than those for IBD), for instance, fatigue may be the dominant contributor to diminished QoL.

The model also did not incorporate adherence and attendance rates for IV iron infusion appointments. As FCM can require more appointments relative to FDI to achieve correction of iron levels (particularly at higher doses) [28,29], it is likely that patients receiving FCM would have lower adherence rates than patients receiving FDI. Hypophosphatemia-related complications (i.e., osteomalacia and subsequent fractures) were also omitted from the model, due to limited publicly-available clinical data. However, the sources used in this study demonstrate that hypophosphatemia rates (particularly severe hypophosphatemia) are significantly higher for patients receiving FCM than patients receiving FDI. If osteomalacia-related QoL and costs were to be included in the model, alongside adherence and attendance rates, it is plausible that FDI may be associated with additional cost savings and further increases QoL relative to FCM.

A key characteristic of this analysis was that the model focused primarily on patients experiencing chronic IDA (i.e., IDA with chronic underlying causes). In this patient group, there is a greater need for multiple, high-dose IV iron doses, which may contribute to the results showing the dominance of FDI over FCM. However, the outcome of dominance for FDI would also hold for acute causes of IDA requiring only one-off or short-term treatment with IV iron (e.g., peri-operative IV iron administration); the proportional clinical, QoL and cost impacts would remain the same for patients with non-chronic acute IDA, simply occurring over a shorter timeframe. The more serious sequelae of chronic hypophosphatemia may not occur in patients with acute IDA, but the omission of these sequelae from the present analyses yields results that are less sensitive to the choice of time horizon.

The analysis was based on relatively conservative estimates of the costs and downstream consequences of hypophosphatemia. Specifically, only those costs associated with monitoring and treatment of hypophosphatemia were considered; any financial implications arising from the complications and sequelae of severe hypophosphatemia (e.g., treatment of hypophosphatemic osteomalacia and fractures) were omitted. The omission was driven by the lack of published data on the fracture rates associated with hypophosphatemia stemming from IV iron administration in China; however, while the omission of costs associated with treating fractures still resulted in an analysis in which FDI was the dominant IV iron formulation, emerging data on fracture incidence after administration of FCM versus FDI suggest that the cost savings with FDI may have been greater still had fractures been incorporated into the analysis [58].

Finally, a patient-level simulation approach was selected as the model design for this analysis, which allowed factors such as individual patient variability, stochastic uncertainty and stochastic variation [32]. This model design allows for increased uncertainty surrounding unique patient-projected outcomes to be incorporated as compared to a cohort-level model design. The use of such a model, alongside the incorporation of parameters representing iron-infusion-related hypophosphatemia and its treatment represent increased levels of robustness, detail and nuance when determining the cost-utility of FDI versus FCM. The model structure used for this analysis therefore serves as a strong foundation for future studies aiming to determine the cost-effectiveness of various IV iron infusions.

## Conclusion

Results showed that FDI would improve patient quality of life and reduce direct healthcare expenditure versus FCM in patients with IDA in China, due to the fact that FDI required comparatively less infusions over five years and had no associated hypophosphatemia-related costs.

## Transparency

### Declaration of funding

Pharmacosmos A/S funded the development of the health economic model and analysis, preparation of the manuscript, and the article processing charge for the manuscript.

### Declaration of financial/other relationships

RFP is a full-time employee, director, and shareholder in, and WA is a full-time employee of, Covalence Research Ltd, which received consultancy fees from Pharmacosmos A/S to develop the patient-level simulation model and analysis and prepare the manuscript.

### Author contributions

RFP conceived of and designed the analysis. RFP developed the simulation model and conducted statistical and health economic analyses. WA conducted health economic and sensitivity analyses, generated tables and figures, and prepared the first draft of the manuscript, which was revised critically for intellectual content by all authors. All authors approved the final version to be published. All authors agree to be accountable for all aspects of the work.

### Previous presentations

Select analyses presented here were presented previously at ISPOR Europe 2023 (Copenhagen, November 12 to November 15, 2023) as poster EE266.

## Data Availability

All data produced in the present study are available upon reasonable request to the authors.

## Notes

### Funding Statement

Pharmacosmos A/S funded the development of the health economic model and analysis, and preparation of the manuscript.

## References

[1] DeLoughery TG. Iron deficiency anemia. Med Clin North Am. 2017;101:319–332.

[2] Kumar A, Sharma E, Marley A, et al. Iron deficiency anaemia: pathophysiology, assessment, practical management. BMJ Open Gastroenterol. 2022;9:e000759.

[3] Pasricha S-R, Tye-Din J, Muckenthaler MU, et al. Iron deficiency. Lancet. 2021;397:233–248.

[4] World Health Organization. Haemoglobin concentrations for the diagnosis of anaemia and assessment of severity [Internet]. Geneva: World Health Organization; 2011 [cited 2021 Sep 10]. Report No.: WHO/NMH/NHD/MNM/11.1. Available from: https://apps.who.int/iris/bitstream/handle/10665/85839/WHO_NMH_NHD_MNM_11.1_eng.pdf?sequence=22&isAllowed=y.

[5] Kassebaum NJ, GBD 2013 Anemia Collaborators. The global burden of anemia. Hematol Oncol Clin North Am. 2016;30:247–308.

[6] Tan J, He G, Qi Y, et al. Prevalence of anemia and iron deficiency anemia in Chinese pregnant women (IRON WOMEN): a national cross-sectional survey. BMC Pregnancy Childbirth. 2020;20:670.

[7] Zheng H, Long W, Tan W, et al. Anaemia, iron deficiency, iron-deficiency anaemia and their associations with obesity among schoolchildren in Guangzhou, China. Public Health Nutr. 2020;23:1693–1702.

[8] Nead KG, Halterman JS, Kaczorowski JM, et al. Overweight children and adolescents: a risk group for iron deficiency. Pediatrics. 2004;114:104–108.

[9] Zhao L, Zhang X, Shen Y, et al. Obesity and iron deficiency: a quantitative meta-analysis. Obes Rev. 2015;16:1081–1093.

[10] Shokrgozar N, Golafshan HA. Molecular perspective of iron uptake, related diseases, and treatments. Blood Res. 2019;54:10–16.

[11] Ando K, Morita S, Higashi T, et al. Health-related quality of life among Japanese women with iron-deficiency anemia. Qual Life Res. 2006;15:1559–1563.

[12] Cooke AG, McCavit TL, Buchanan GR, et al. Iron deficiency anemia in adolescents who present with heavy menstrual bleeding. J Pediatr Adolesc Gynecol. 2017;30:247– 250.

[13] Mirza FG, Abdul-Kadir R, Breymann C, et al. Impact and management of iron deficiency and iron deficiency anemia in women’s health. Expert Rev Hematol. 2018;11:727–736.

[14] Anker SD, Kirwan B-A, van Veldhuisen DJ, et al. Effects of ferric carboxymaltose on hospitalisations and mortality rates in iron-deficient heart failure patients: an individual patient data meta-analysis. Eur J Heart Fail. 2018;20:125–133.

[15] Danese S, Hoffman C, Vel S, et al. Anaemia from a patient perspective in inflammatory bowel disease: results from the European Federation of Crohn’s and Ulcerative Colitis Association’s online survey. Eur J Gastroenterol Hepatol. 2014;26:1385.

[16] Gisbert JP, Bermejo F, Pajares R, et al. Oral and intravenous iron treatment in inflammatory bowel disease: hematological response and quality of life improvement. Inflamm Bowel Dis. 2009;15:1485–1491.

[17] García-López S, Bocos JM, Gisbert JP, et al. High-dose intravenous treatment in iron deficiency anaemia in inflammatory bowel disease: early efficacy and impact on quality of life. Blood Transfus. 2016;14:199–205.

[18] Taylor S, Rampton D. Treatment of iron deficiency anemia: practical considerations. Pol Arch Med Wewn. 2015;125:452–460.

[19] Onken JE, Bregman DB, Harrington RA, et al. A multicenter, randomized, active-controlled study to investigate the efficacy and safety of intravenous ferric carboxymaltose in patients with iron deficiency anemia. Transfusion (Paris). 2014;54:306–315.

[20] Rozen-Zvi B, Gafter-Gvili A, Paul M, et al. Intravenous versus oral iron supplementation for the treatment of anemia in CKD: systematic review and meta-analysis. Am J Kidney Dis. 2008;52:897–906.

[21] Koutroubakis IE, Oustamanolakis P, Karakoidas C, et al. Safety and efficacy of total-dose infusion of low molecular weight iron dextran for iron deficiency anemia in patients with inflammatory bowel disease. Dig Dis Sci. 2010;55:2327–2331.

[22] Schröder O, Mickisch O, Seidler U, et al. Intravenous iron sucrose versus oral iron supplementation for the treatment of iron deficiency anemia in patients with inflammatory bowel disease--a randomized, controlled, open-label, multicenter study. Am J Gastroenterol. 2005;100:2503–2509.

[23] Boots JMM, Quax RAM. High-dose intravenous iron with either ferric carboxymaltose or ferric derisomaltose: a benefit-risk assessment. Drug Saf. 2022;45:1019–1036.

[24] Schaefer B, Tobiasch M, Wagner S, et al. Hypophosphatemia after intravenous iron therapy: comprehensive review of clinical findings and recommendations for management. Bone. 2022;154:116202.

[25] Zoller H, Schaefer B, Glodny B. Iron-induced hypophosphatemia: an emerging complication. Curr Opin Nephrol Hypertens. 2017;26:266–275.

[26] Glaspy JA, Lim-Watson MZ, Libre MA, et al. Hypophosphatemia associated with intravenous iron therapies for iron deficiency anemia: a systematic literature review. Ther Clin Risk Manag. 2020;16:245–259.

[27] Wolf M, Rubin J, Achebe M, et al. Effects of iron isomaltoside vs ferric carboxymaltose on hypophosphatemia in iron-deficiency anemia: two randomized clinical trials. JAMA. 2020;323:432–443.

[28] emc. Ferinject (ferric carboxymaltose) – Summary of Product Characteristics (SmPC) [Internet]. [cited 2022 Oct 18]. Available from: https://www.medicines.org.uk/emc/product/5910/smpc.

[29] emc. Ferric derisomaltose Pharmacosmos 100 mg/ml solution for injection/infusion – Summary of Product Characteristics (SmPC) [Internet]. [cited 2023 Jul 12]. Available from: https://www.medicines.org.uk/emc/product/5676/smpc.

[30] Greenhawt M, Oppenheimer J, Codispoti CD. A practical guide to understanding cost-effectiveness analyses. J Allergy Clin Immunol Pract. 2021;9:4200–4207.

[31] Hu S, Liu L, Pollock RF, et al. Intravenous iron for the treatment of iron deficiency anemia in China: a patient-level simulation model and cost-utility analysis comparing ferric derisomaltose with iron sucrose. J Med Econ. 2022;25:561–570.

[32] Davis S, Stevenson M, Tappenden P, et al. NICE DSU Technical Support Document 15: Cost-Effectiveness Modelling Using Patient-Level Simulation. Sheffield; 2014.

[33] Zoller H, Wolf M, Blumenstein I, et al. Hypophosphataemia following ferric derisomaltose and ferric carboxymaltose in patients with iron deficiency anaemia due to inflammatory bowel disease (PHOSPHARE-IBD): a randomised clinical trial. Gut. 2023;72:644–653.

[34] Pollock RF, Muduma G. A systematic literature review and indirect comparison of iron isomaltoside and ferric carboxymaltose in iron deficiency anemia after failure or intolerance of oral iron treatment. Expert Rev Hematol. 2019;12:129–136.

[35] Bellos I, Frountzas M, Pergialiotis V. Comparative risk of hypophosphatemia following the administration of intravenous iron formulations: a network meta-analysis. Transfus Med Rev. 2020;34:188–194.

[36] Blumenstein I, Shanbhag S, Langguth P, et al. Newer formulations of intravenous iron: a review of their chemistry and key safety aspects – hypersensitivity, hypophosphatemia, and cardiovascular safety. Expert Opin Drug Saf. 2021;20:757– 769.

[37] Vifor International. To assess the impact of ferric carboxymaltose compared with iron sucrose in Chinese subjects on correcting iron deficiency anaemia: study results [Internet]. Vifor International; 2021 [cited 2023 Jul 10]. Available from: https://clinicaltrials.gov/ct2/show/results/NCT03591406.

[38] Kulnigg S, Teischinger L, Dejaco C, et al. Rapid recurrence of IBD-associated anemia and iron deficiency after intravenous iron sucrose and erythropoietin treatment. Am J Gastroenterol. 2009;104:1460–1467.

[39] Chen Y, Guan H, Han S, et al. China guidelines for pharmacoeconomic evaluations: 2020 edition [Internet]. Chinese Pharmaceutical Association and China Society for Pharmacoeconomics and Outcomes Research and ISPOR Beijing Chapter; 2020 [cited 2022 Jan 4]. Available from: https://tools.ispor.org/PEguidelines/source/China-Guidelines-for-Pharmacoeconomic-Evaluations-2020.pdf.

[39] National Bureau of Statistics of China. China Statistical Yearbook 2022 [Internet]. [cited 2023 Jul 11]. Available from: http://www.stats.gov.cn/sj/ndsj/2022/indexeh.htm.

[41] Kassianides X, Bhandari S. Hypophosphataemia, fibroblast growth factor 23 and third-generation intravenous iron compounds: a narrative review. Drugs Context. 2021;10:2020-11–13.

[42] Medicines and Healthcare products Regulatory Agency. Drug safety update: ferric carboxymaltose (Ferinject▾): risk of symptomatic hypophosphataemia leading to osteomalacia and fractures [Internet]. GOV.UK. 2020 [cited 2021 Aug 12]. Available from: https://www.gov.uk/drug-safety-update/ferric-carboxymaltose-ferinject-risk-of-symptomatic-hypophosphataemia-leading-to-osteomalacia-and-fractures.

[43] Vifor (International) Inc. 羧基麦芽糖铁注射液说明书 (Ferric Carboxymaltose Injection Instructions). 2022.

[44] Fresenius Kabi China. 格利福斯® – 费森尤斯卡比中国 [Internet]. [cited 2024 May 11]. Available from: https://www.fresenius-kabi.com/cn/我们的产品/格利福斯.

[45] Pharmacosmos A/S. A trial comparing the incidence of hypophosphatemia in relation to treatment with iron isomaltoside and ferric carboxymaltose in subjects with iron deficiency anaemia due to inflammatory bowel disease [Internet]. 2021 [cited 2023 Jul 11]. Available from: https://clinicaltrials.gov/ct2/show/NCT03466983.

[46] Brennan VK, Dixon S. Incorporating process utility into quality adjusted life years: a systematic review of empirical studies. PharmacoEconomics. 2013;31:677–691.

[47] Hu S, Wu D, Wu J, et al. Disutilities associated with intravenous iron infusions: results from a time trade-off survey and diminishing marginal utility model for treatment attributes in China. Patient Relat Outcome Meas. 2023;14:253–267.

[48] Kangzhou Big Data (Group) Co., Ltd. 健康产业大数据服务与赋能平台: Ferric Derisomaltose [Internet]. [cited 2024 May 11]. Available from: https://www.yaozh.com/.

[49] Kangzhou Big Data (Group) Co., Ltd. 健康产业大数据服务与赋能平台: Ferric Carboxymaltose [Internet]. [cited 2024 May 11]. Available from: https://www.yaozh.com/.

[50] 博鳌恒大国际医院 (Boao Evergrande International Hospital). Public procurement platform of Hainan Boao Evergrande International Hospital [Internet]. [cited 2024 May 11]. Available from: http://boaohospital.com/.

[51] Kangzhou Big Data (Group) Co., Ltd. 健康产业大数据服务与赋能平台: Sodium Glycerophosphate Injection [Internet]. [cited 2024 May 11]. Available from: https://www.yaozh.com/.

[52] Pollock RF, Kalra PA, Kalra PR, et al. A systematic review, meta-analysis, and indirect comparison of blindly adjudicated cardiovascular event incidence with ferric derisomaltose, ferric carboxymaltose, and iron sucrose. Adv Ther. 2022;39:4678– 4691.

[53] Pollock RF, Biggar P. Indirect methods of comparison of the safety of ferric derisomaltose, iron sucrose and ferric carboxymaltose in the treatment of iron deficiency anemia. Expert Rev Hematol. 2020;13:187–195.

[54] Pollock RF, Muduma G. An economic analysis of ferric derisomaltose versus ferric carboxymaltose in the treatment of iron deficiency anemia in patients with inflammatory bowel disease in Norway, Sweden, and Finland. Clin Outcomes Res. 2021;Volume 13:9–18.

[55] Pollock RF, Muduma G. A patient-level cost-effectiveness analysis of iron isomaltoside versus ferric carboxymaltose for the treatment of iron deficiency anemia in the United Kingdom. J Med Econ. 2020;23:751–759.

[56] Nocerino A, Nguyen A, Agrawal M, et al. Fatigue in inflammatory bowel diseases: etiologies and management. Adv Ther. 2020;37:97–112.

[57] McDowell C, Farooq U, Haseeb M. Inflammatory bowel disease. StatPearls [Internet]. Treasure Island (FL): StatPearls Publishing; 2023 [cited 2023 Dec 15]. Available from: http://www.ncbi.nlm.nih.gov/books/NBK470312/.

[58] Zoller H, Pammer LM, Schaefer B, et al. Incidence of fractures after intravenous iron: a retrospective analysis comparing ferric carboxymaltose and ferric derisomaltose. Blood [Internet]. ASH; 2023 [cited 2023 Nov 10]. p. 3838. Available from: https://ash.confex.com/ash/2023/webprogram/Paper174508.html.

